# Slower reactive stepping kinematics are associated with lower clinical balance function and delayed cortical evoked responses under challenging balance conditions after stroke

**DOI:** 10.64898/2025.12.18.25342582

**Authors:** Si-Yu Tsai, Aiden M Payne, Jasmine L Mirdamadi, Lena H Ting, Michael R Borich, Jacqueline A Palmer

## Abstract

**Background:** The ability to elicit a rapid, reactive step to recover balance after a postural destabilization is paramount to fall prevention. In response to a given balance perturbation magnitude, people after stroke display impaired spatiotemporal stepping kinematics. Yet, spatiotemporal stepping kinematics at individualized perturbation magnitudes after stroke and the underlying neural correlates remain unknown. Here, we tested whether stepping kinematics differ in people after stroke at individualized balance perturbation magnitudes and further examined neuromechanical mechanisms underlying impaired stepping kinematics after stroke.

**Methods:** 16 participants with chronic (>6mo.) stroke and 16 age-matched controls underwent standing balance perturbations at individualized step threshold, the perturbation magnitude that elicited unintentional steps in approximately 50% of trials. We quantified the spatiotemporal kinematics of the first reactive step and weight-bearing asymmetry immediately prior to the perturbation onset. Cortical N1s, perturbation-evoked brain responses reflecting cortical processing for balance control, were extracted from Cz signals and identified as the most negative local minimum (100-300ms).

**Results:** While there were no group-level differences in step duration, step velocity, and step trajectory, people with stroke showed delayed step initiation (Cohen’s d= 0.89, p=0.02) and termination latencies (Cohen’s d= 0.81, p=0.03). Delayed step initiation latencies after stroke correlated with lower clinical balance function (e.g., miniBEST score; r= -0.67, p=0.004) and delayed cortical responses (r=0.58, p=0.02) but not weight-bearing asymmetry (p>0.86).

**Conclusions:** The relationships between delayed step initiation, lower clinical balance function, and slower cortical responses suggests cortical processing speed may be a limiting factor for post-stroke balance function and identifies neuromechanical targets for fall prevention.

## Introduction

The initiation of a rapid reactive step is crucial for balance recovery and fall prevention.^1–5^ While people after stroke display less effective stepping kinematics, including decreased step length,^5–7^ slower step velocity,^6^ and delayed step initiation,^3,5^ that are associated with higher fall incidence,^2,3^ one limitation in the current literature is that perturbations are delivered at a fixed magnitude across individuals.^3,5,8^ Given well-characterized individual variability in balance ability, the same perturbation magnitude may differ in challenge levels across individuals, which may explain differences in reactive balance recovery. This introduces confounds when comparing reactive stepping kinematics across individuals with different balance abilities, such as people with and without post-stroke balance impairment, as stepping kinematics vary as a function of balance difficulty.^3,6^ Comparing reactive stepping kinematics between groups while matching task difficulty across individuals allow us to assess stroke-specific effects without the confounds introduced by a mismatch in task difficulty.

Reactive stepping responses require one leg to initiate a reactive step while the stance leg supports body weight.^7^ When not explicitly constrained, people with stroke generally prefer to step with their non-paretic leg,^1,4,9^ which forces them to shift their weight onto the paretic leg in order to unload their non-paretic leg to take a step.^3,4^ It is therefore not surprising that weight-bearing asymmetry contributes to delayed step initiation that are initiated by the non-paretic leg, as people with stroke tend to bear more weight on their non-paretic leg and thus have to unload the weight to take a step.^3,10^ Further, only delayed step initiation latencies of the non-paretic leg are associated with higher odds of perturbation-evoked falls.^3^ Yet, reactive stepping behavior and its link to weight-bearing asymmetry after stroke at individualized perturbation intensities remain unclear.

During balance recovery, an evoked cortical response occurs on the timescale that is early enough to allow for the possibility of cortical influences on reactive stepping responses.^11–13^ The cortical N1 response is a perturbation-evoked brain response that has been localized to the supplementary motor area^14–16^ and is believed to reflect cortical processing of balance error^17^ and/or postural threats.^18^ One recent study assessing how cortical N1 responses change as a function of perturbation intensity and response strategy (e.g., feet-in-place and stepping responses) showed that the cortical N1 scales with perturbation intensity and relates to reactive stepping behavior in younger adults.^13,19^ Our group recently reported that delayed cortical N1 responses after stroke are associated with clinical balance dysfunction and delayed reactive balance kinetics during feet-in-place recovery responses.^20^ Yet, it is unknown whether the atypical cortical N1 response is related to impaired reactive stepping kinematics after stroke, particularly under individualized, challenging balance conditions.

In the present study, we analyzed data from the session where we scaled up perturbation magnitudes to an individualized balance challenge point that elicited reactive stepping responses, and assessed stroke-specific neuromechanical characteristics of reactive stepping responses at comparable challenging balance perturbations.^20^ The purpose of this study was to 1) compare spatiotemporal stepping kinematics at individualized balance challenge points in older adults with and without stroke and 2) test the relationships between reactive stepping kinematics, balance function, and weightbearing asymmetry. Additionally, as an exploratory analysis, we assessed the relationship between cortical evoked responses and reactive stepping kinematics. Characterizing impaired reactive stepping kinematics after stroke and identifying the underlying neuromechanical factors could inform fall-preventing rehabilitation strategies.

## Methods

### Participants and study procedures

This secondary analysis builds on biomechanics and electroencephalography (EEG) measures of cortical activity during feet-in-place responses to smaller balance perturbations that have been previously reported in this cohort.^20^ Sixteen individuals with chronic (>6 months) stroke and 16 age-matched controls completed a single visit of reactive balance, and clinical testing (i.e., Mini Balance Evaluation Systems Test (miniBEST), Timed-Up-and-Go (TUG), and 10-m walk test). Inclusion criteria included age>21 years old, the ability to walk at least 10 m without the assistance of another person, the ability to stand unassisted for at least 3 minutes, and the cognitive ability to provide informed consent. Participants were excluded if they had any diagnosed neurologic condition other than stroke, or pain affecting standing or walking. Clinical characteristics of this cohort can be found in Table 1 and stroke lesion characteristics for this cohort are described in Palmer et al. (2024).^20^ The experimental protocol was approved by the Emory University Institutional Review Board, and all participants provided written informed consent.

**Table 1.**
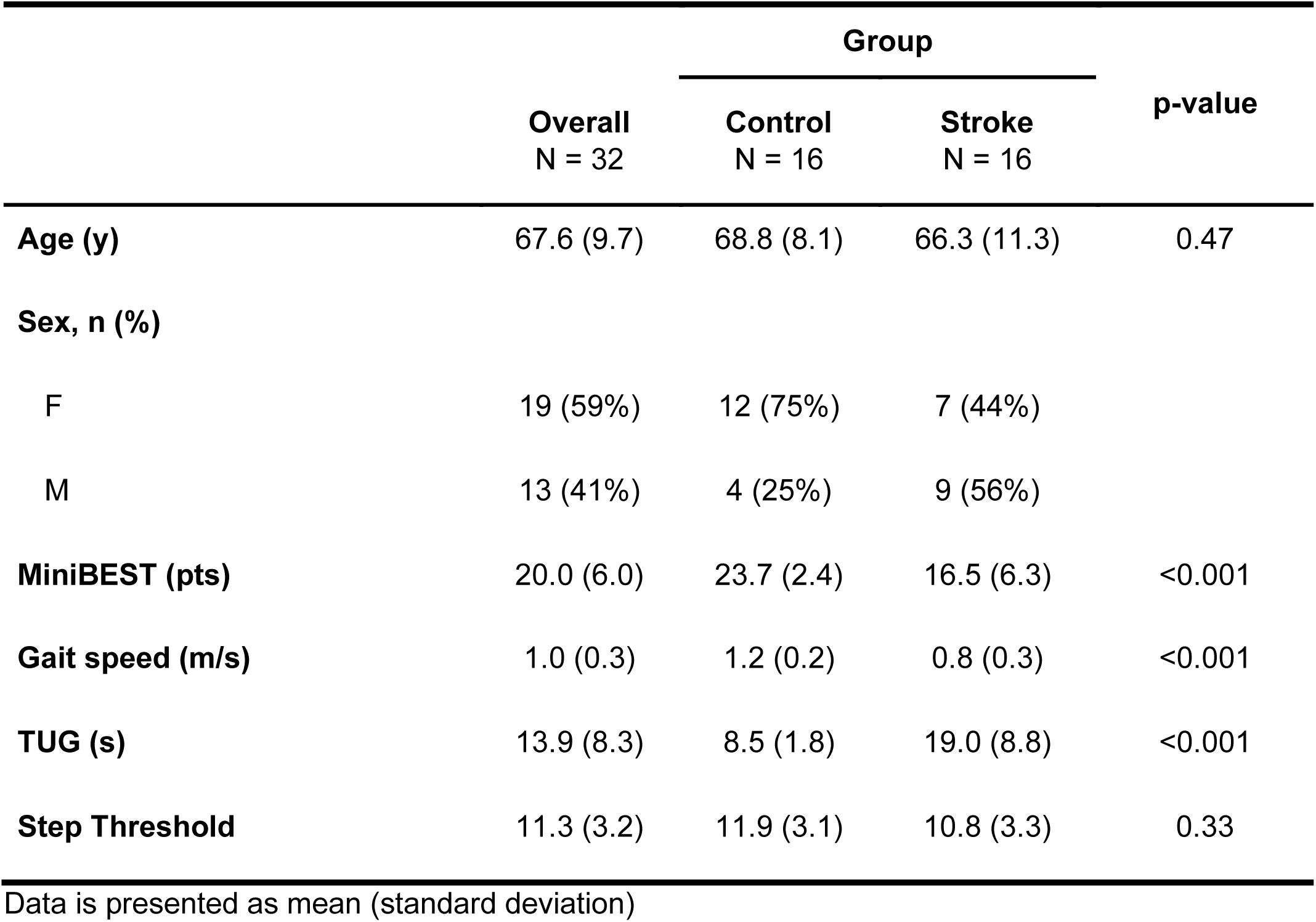
Participant characteristics.

#### Standing Balance Perturbations

Participants stood barefoot in their typical, self-selected posture with feet approximately shoulder width apart, and arms folded across their chest on a moving platform (Factory Automation Systems, Atlanta, GA) and received support-surface translational perturbations. Participants were instructed to recover balance with a feet-in-place strategy. Baseline stance width was not precisely standardized as similar motor response latencies are observed across a range of narrow and wider stances.^21^ Participants first underwent a series of multidirectional perturbations at a standardized, lower perturbation magnitude, intended to elicit feet-in-place responses, as reported previously.^20^ After a seated rest break, participants then underwent a second series of anteriorly-directed perturbations to determine their individualized step threshold. Step threshold was determined as the perturbation magnitude that elicited unintentional reactive stepping responses in approximately 50% of trials.^22^ Posteriorly-directed perturbations were delivered once approximately every 10 trials to prevent participants from anticipating anteriorly-directed perturbations. Only perturbations delivered at the individual’s reactive step threshold and that elicited a reactive step to recover balance were included in the present analyses, and have not been reported previously. Participants took a seated rest break every 8 minutes, or more frequently if requested or signs of fatigue emerged. Experimenters closely monitored real-time kinetics between trials to ensure that participants maintained an upright, vertical body position in the sagittal plane. If real-time forces in either limb deviated from the vertical plane, participants were verbally corrected for postural lean and cued to stand upright. For trials that elicited reactive steps, the experimenter ensured that feet placement was returned to baseline prior to the start of the next trial.

### Kinetic and Kinematic Data Acquisition and Analysis

Kinetic (2 AMTI OR6-6 force plates, each 40 cm × 60 cm; sampling rate 1000Hz) and kinematic (10-camera Vicon Nexus 3D motion analysis system; sampling rate 100Hz) data were recorded during balance recovery. 42 reflective markers were placed on anatomical landmarks on the legs and trunk, in accordance with the modified Helen-Hayes set.

#### Spatiotemporal metrics of stepping responses

The raw data were first gap-filled then post-processed using the Vicon plug-in gait model pipeline. The output data were then low-passed filtered using a fourth-order Butterworth filter with a cut-off frequency of 6 Hz.^5,6^

Spatiotemporal metrics of the first step were computed from the heel marker Z-coordinate (i.e., vertical axis) trajectory. Step initiation latency was identified as the time at which the Z-coordinate exceeded 2 standard deviations above the baseline, while step termination latency was identified as the time at which the Z-coordinate returned to 2 standard deviations above the baseline.^5,6^ In some trials, participants failed to elevate their foot to a height that could reliably be used to identify step initiation; for these trials, the relative displacement in the Y-coordinate (i.e., anteroposterior axis) between the two heel markers was used to identify step initiation using the same criteria. Step termination was identified as the time at which the displacement of the stepping foot relative to the stance foot reached its first local maximum in the anteroposterior direction. The step duration was then calculated as the time between step initiation and termination. Step length and step height were defined as the maximal relative displacement between the two heel markers in the anteroposterior direction and absolute vertical displacement of the stepping foot, respectively. Average step velocity was calculated as the average velocity over the step duration in the anteroposterior axis. All the kinematic variables were verified through visual inspection. All the variables were computed using a customized script in MATLAB software (The MathWorks Inc.)

#### Weight-bearing asymmetry index

Weight-bearing asymmetry index was computed from the mean of the vertical force vector for each stepping trial 1s prior to the perturbation onset between two force plates and was quantified as:

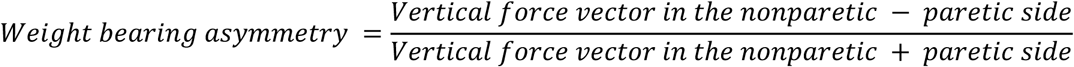

The right leg was matched to the non-paretic side, while the left leg was matched to the paretic side for controls.^22^ A positive value indicated that individuals bear more weight on the non-paretic leg (stroke group) or right leg (control group) immediately prior to the perturbation onset. We additionally analyzed the absolute value of the index to examine whether the degree of weight-bearing asymmetry differs between groups and the relationship to stepping kinematics.

### EEG Data Acquisition and Analysis

Cortical activity during standing balance perturbations was continuously recorded (sampling rate 1000Hz) using EEG, with a 64-channel active electrode cap and ActiCHamp amplifier with a 24-bit A/D converter and an online 20 kHz anti-aliasing low-pass filter (Brain Products, GmbH). The raw EEG data were preprocessed in EEGlab using the BeMoBIL Pipeline.^23^ BeMoBIL preprocessing consisted of line noise removal using the Zapline plus plugin and bad channel detection and removal using the cleanrawdata plugin. Bad channels were interpolated and then full-rank average referenced. Preprocessed data were then high-pass filtered with a cut-off of 1.75 Hz and epoched -2 to 2 seconds around each perturbation, given the discrete nature of the task.^24^ Maximally independent components (ICs) were extracted using the adaptive mixture component analysis algorithm (AMICA).^25^ ICs that were identified by the algorithm as non-brain sources were further verified through visual inspection before removal. Since AMICA and evoked response analyses require different filtering, we then reverted to the BeMoBIL preprocessed unfiltered continuous data to apply different filters before using the identified components to remove non-brain sources. Specifically, the unfiltered data were low-pass filtered with a cutoff of 30 Hz and high-pass filtered with a cutoff of 0.5Hz, epoched -2 to 2 seconds, reconstructed with the identified components, excluding the non-brain sources, reverted back to channel space, and baseline subtracted -150 to - 50 ms before perturbation onset.^24,26^ The cortical N1 response over the midline sensorimotor region (Cz) was then identified as the most negative local minimum point in the mean Cz EEG waveform between 100-300ms post-perturbation across stepping trials at the magnitude of the participant’s step threshold.^20^

### Statistical analysis

Normality and heterogeneity of variances were verified using the Shapiro-Wilk test and Levene’s test, respectively. Independent t-tests were conducted to examine group-level differences on clinical metrics, step threshold, weight-bearing asymmetry index, and spatiotemporal stepping kinematics. To further examine factors underlying impaired reactive stepping kinematics after stroke, Pearson’s correlation analyses were performed to investigate the relationships between reactive stepping kinematics and clinical, weightbearing asymmetry, and cortical N1 peak metrics within each group. All analyses were performed using R software version 4.4.2^27^ with a priori level of significance set to 0.05.

## Results

People with stroke had lower miniBEST scores (Cohen’s d=1.49), slower gait speed (Cohen’s d=1.68), and slower TUG performance (Cohen’s d=1.62) compared to the age-matched control group. Technical issues with biomechanical data acquisition occurred in 3 participants (stroke, n = 2; control, n = 1), who were subsequently excluded from analyses.

### Reactive stepping biomechanical and behavioral performance metrics

Fifteen out of 16 participants in the stroke group preferred to take a reactive step with their nonparetic leg **(Figure 1).** Reactive step thresholds were not different between stroke and age-matched controls (p=0.33) **(Figure 2A)**. People with stroke had longer step initiation (stroke=0.58±0.19s, controls=0.44±0.09s, t=-2,52, p=0.02) and termination latencies (stroke=0.85±0.2s, controls=0.71±0.1s, t=-2.31, p=0.03) compared to age-matched controls **(Figure 2B)**. Yet, there was no group-level difference in step duration **(Figure 2B)**. No group-level differences were observed in stepping kinematics measured as step length (p= 0.59), step height (p=0.42), or step velocity (p=0.61) **(Figure 2C)**.

**Figure 1.**
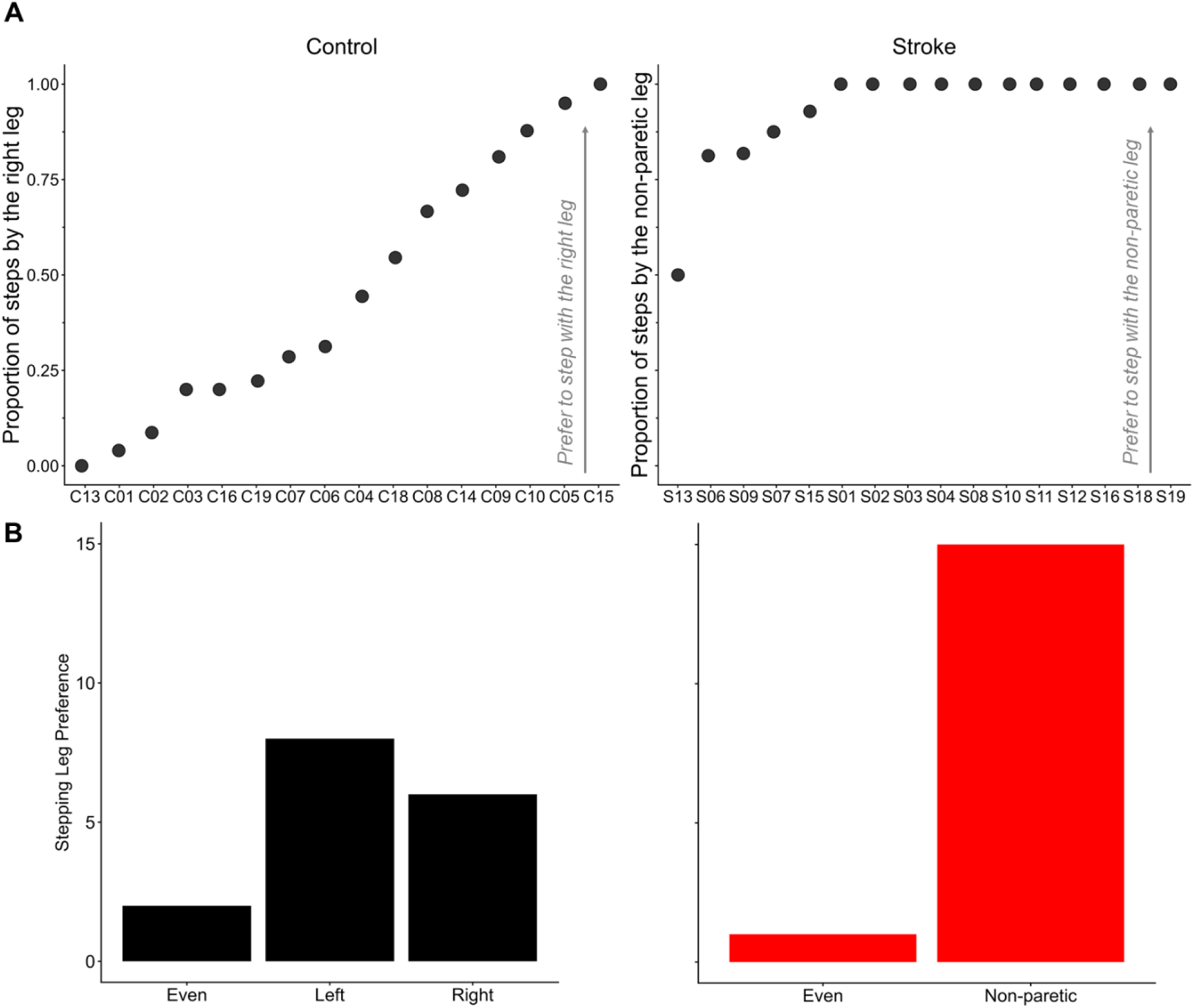
(A) Stepping leg distribution and (B) preference across people after stroke and controls. While controls as a group showed no clear preferred stepping leg, the majority of participants in stroke group (15 out of 16) preferred to initiate the first reactive step with their non-paretic leg. Stepping leg preference was determined as the leg with the greatest proportion of first steps that met or exceeded 70% of all stepping trials.

**Figure 2.**
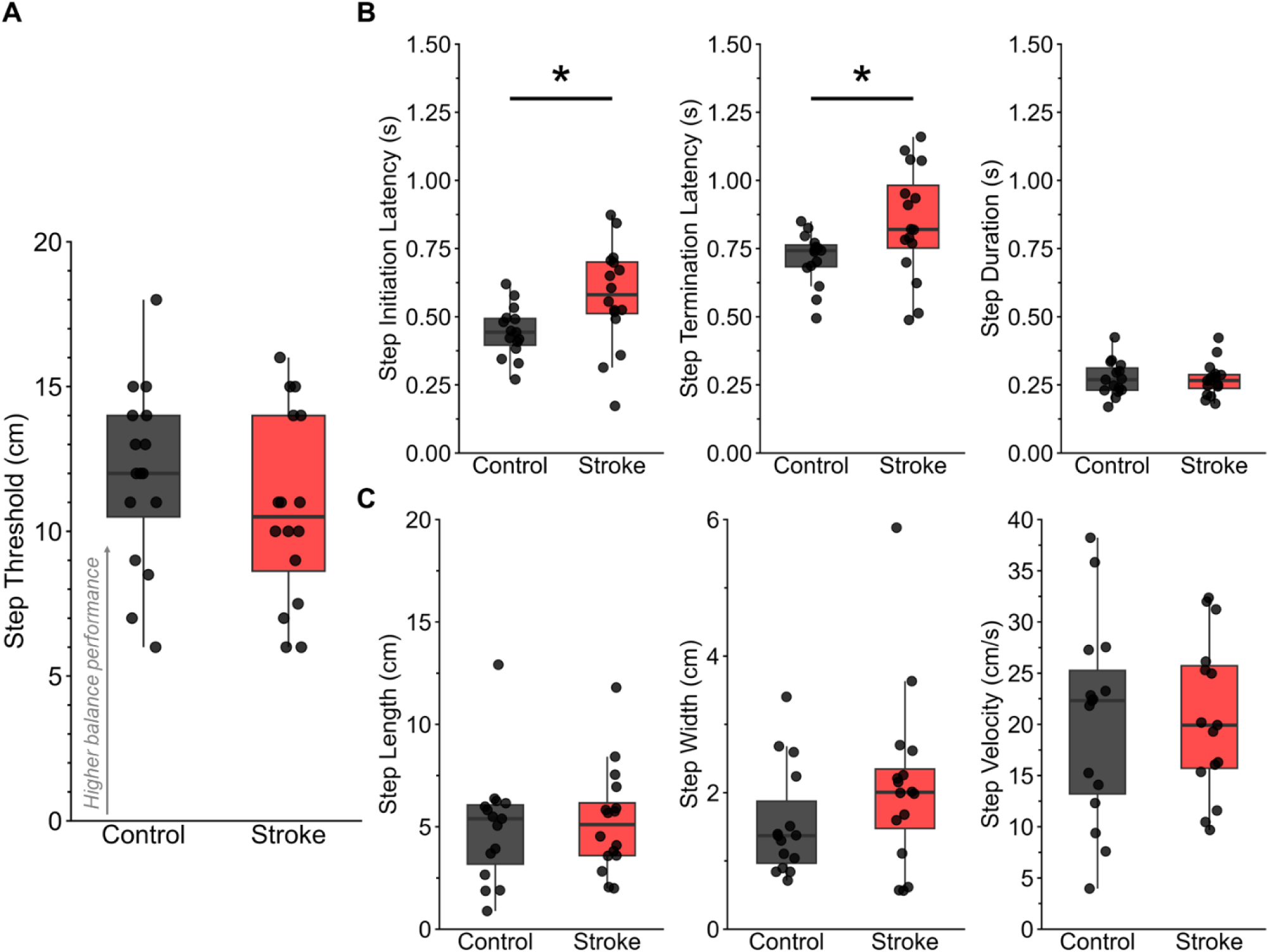
Balance perturbation magnitude and spatiotemporal stepping kinematics at each individual’s reactive step threshold. **(A)** The perturbation magnitude at reactive step threshold was not statistically different in people with stroke compared to age-matched controls, among high between-subject variability (p=0.33). **(B)** People after stroke showed delayed step initiation (p=0.02) and termination latencies (p=0.03) compared to age-matched controls. However, there was no group-level difference in total step duration (p=0.86). **(C)** There were no group-level differences in either step length, step height, or average step velocity (all p>0.42).

### Relationships between step initiation latencies and clinical balance and walking metrics

Delayed step initiation latencies after stroke were associated with lower miniBEST scores (r=-0.67, p=0.004) **(Figure 3A),** but not TUG **(Figure 3B)** or gait speed **(Figure 3C)**. There was no between-group difference in weight bearing asymmetry index (controls: 0.03±0.06, range: [-0.08, 0.17]; stroke: -0.01±0.12, range: [-0.18, 0.3] (p=0.47)) or the absolute value of asymmetry (p=0.12) **(Figure 4A)**, and no relationships were observed between delayed step initiation latencies and weight-bearing asymmetry index in stroke or controls (all p>0.86) **(Figure 4B)**. At the group-level, delayed step initiation latencies were associated with delayed N1 peak latency in people with stroke (r=0.58, p=0.02) (**Figure 5B**). There was no significant relationship between N1 peak amplitude and delayed step initiation in stroke or controls (all p>0.48). Delayed N1 latency was associated with lower miniBEST scores in stroke (r=-0.79, p<0.01) but not in controls (r=-0.17, p=0.54), similar to our previous report in this cohort during smaller balance perturbations.^20^

**Figure 3.**
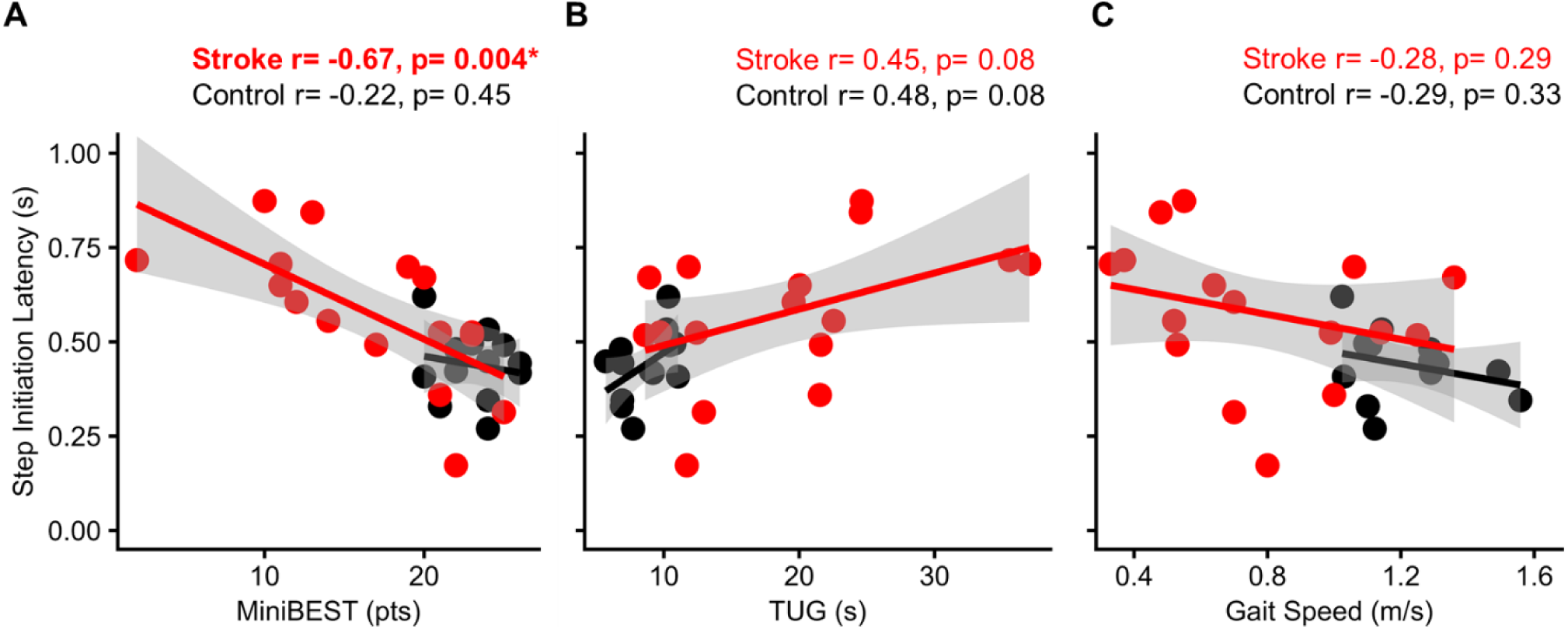
Relationships between step initiation latencies and clinical balance and walking metrics. In stroke, more delayed step initiation latency associated with lower **(A)** miniBEST score, yet showed no relationship with **(B)** TUG performance and **(C)** gait speed. No relationships were observed in controls between step initiation and any clinical balance or walking metric assessed.

**Figure 4.**
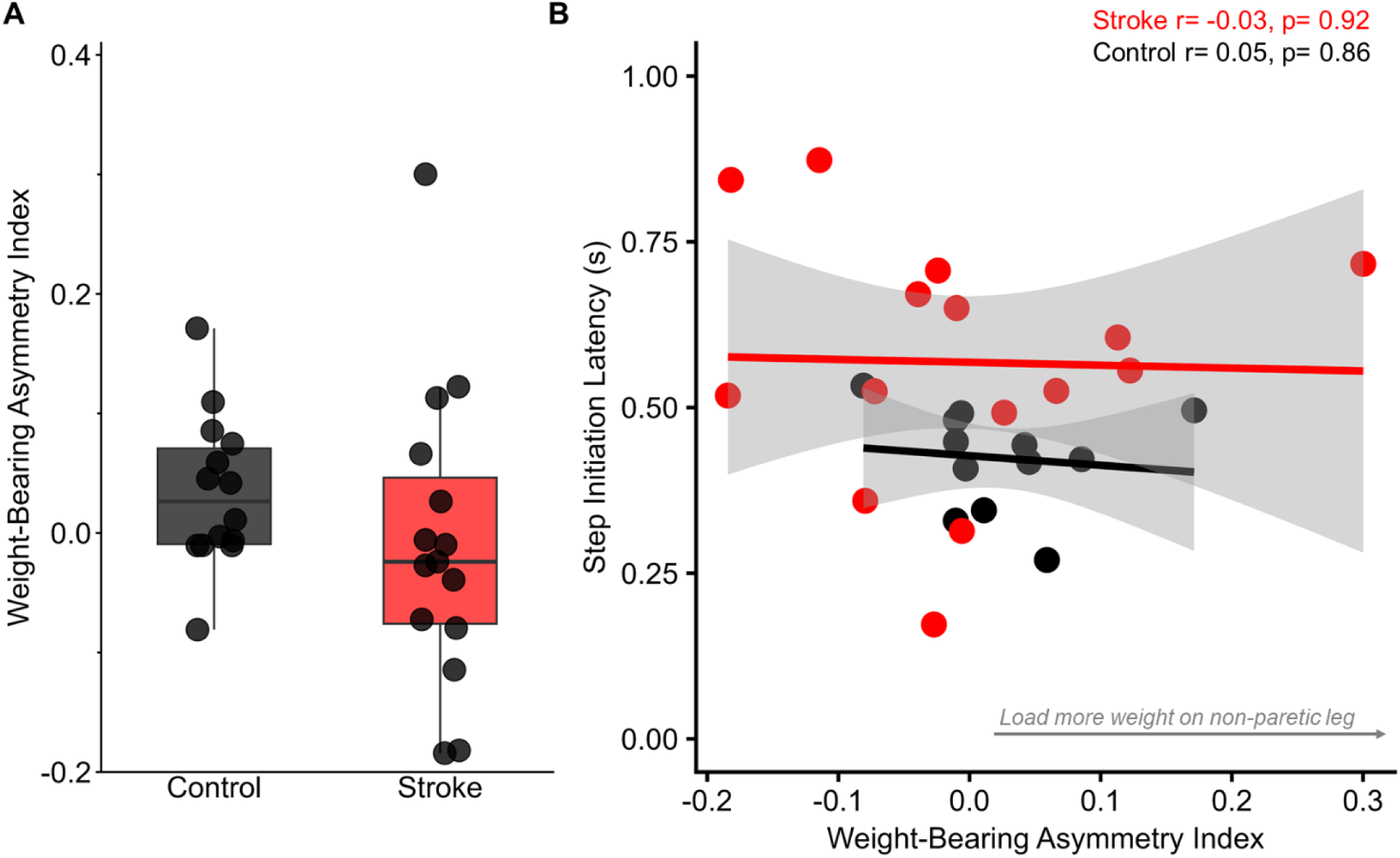
Weight-bearing asymmetry index and relationships to stepping kinematics. **(A)** There was no difference in weight-bearing asymmetry index during baseline standing prior to the perturbation onset between stroke and control groups (p=0.47). **(B)** There were no relationships between weight-bearing asymmetry index and step initiation latencies across stroke and control groups.

**Figure 5.**
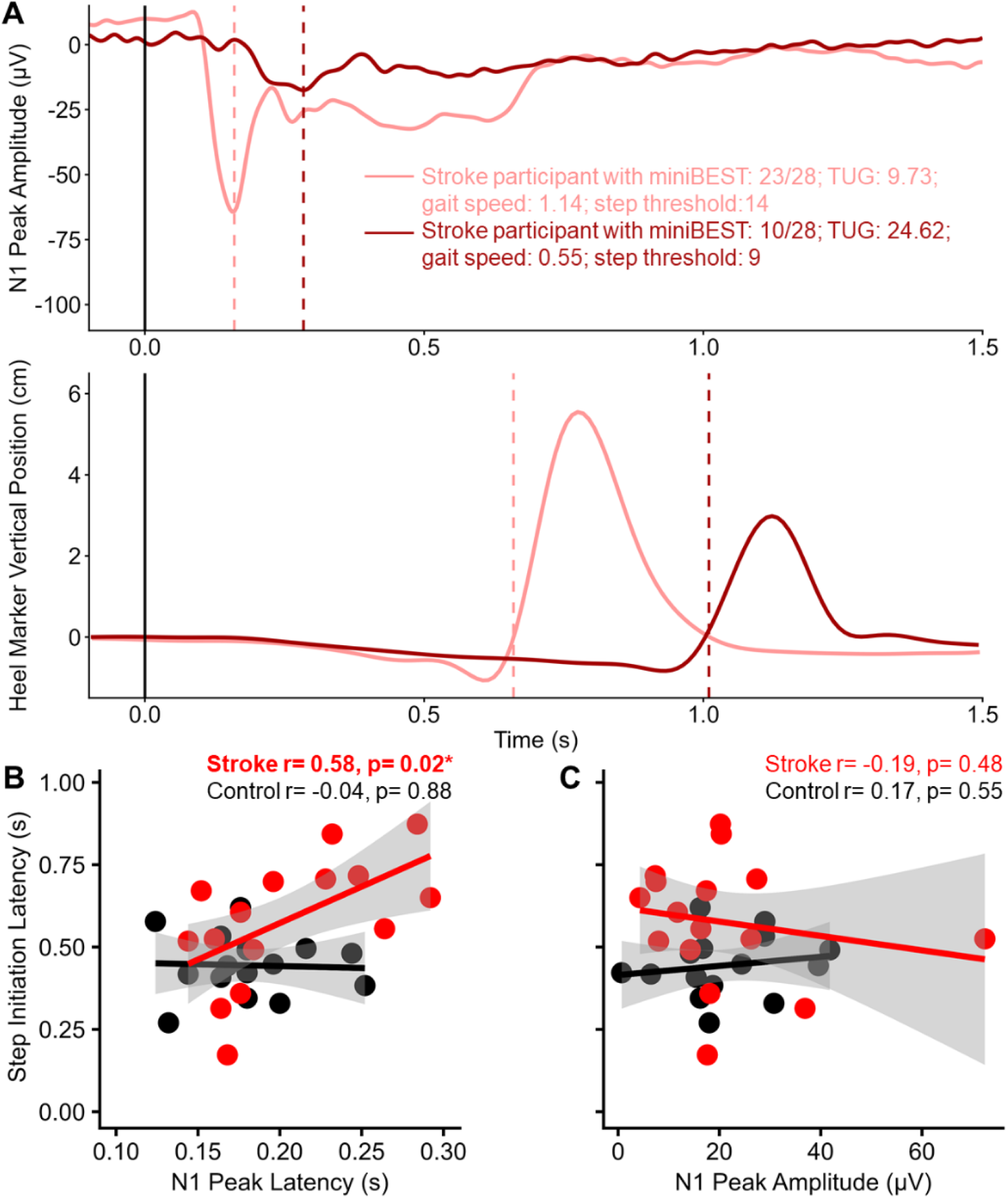
Exemplar cortical N1 responses, reactive stepping kinematics, and relationships across groups. **(A)** Mean N1 waveforms (top) and single-trial heel marker vertical trajectories (bottom). individuals with higher and lower balance function in stroke group. The perturbation onset was denoted at time=0 and color-coded as black solid lines, while the latencies of cortical N1 responses and step initiation were illustrated as dash-lines. An exemplar individual with higher balance function (miniBEST: 23/28; TUG: 9.73; gait speed: 1.14m/s; step threshold: 14) showed faster N1 responses and step initiation (maroon). An exemplar individual with lower balance function (miniBEST: 10/28; TUG: 24.62; gait speed: 0.55; step threshold: 9) had slower N1 responses and step initiation (pink). **(B)** The delayed step initiation after stroke correlates with delayed cortical N1 responses, while showing no relationships in controls **(C)** Neither stroke nor controls showed relationships between step initiation and N1 peak amplitude.

## Discussion

Our findings show that the ability to initiate rapid, reactive steps under challenging balance conditions is a key balance impairment that may limit clinical balance function in people after stroke. The relationships between delayed cortical N1 responses and delayed step initiation in stroke implicates the speed of cortical processing as a potential neural substrate underpinning the limits of reactive step initiation after stroke.

The relationship between delayed step initiation and lower balance ability after stroke suggests that delayed step initiation may contribute to clinical balance dysfunction after stroke. This finding aligns with a previous finding that delayed step initiation by the non-paretic leg is associated with fall incidence after stroke.^3^ Contrary to previous findings,^3^ delayed step initiation after stroke was not explained by weight-bearing asymmetry in the present study. The discrepancy could be explained by differences in experimental paradigms (i.e., lean-and-release perturbation paradigms that elicit forward reactive steps^3^ compared to support-surface perturbations that elicit backward reactive steps in the present study). Different perturbation paradigms might probe distinct neuromechanical mechanisms for reactive balance recovery.^28^

### Delayed cortical processing may contribute to delayed step initiation latencies after stroke under challenging balance conditions

People with stroke appear to increase reliance on cortical resources to compensate for impaired balance and mobility, as indicated by greater cognitive dual-task interference during locomotion^29^ and positive relationships between cortical activity during balance recovery and clinical balance function.^20,30^ While recruitment of cortical resources may enable people after stroke to compensate for motor impairment at lower levels of balance difficulty, this neural strategy may impose a “ceiling” for neuromotor control under challenging balance conditions.^31,32^ Our previous findings showed that people after stroke have smaller and delayed cortical activity evoked during balance recovery to smaller perturbations at a standardized magnitude that may reflect their inability to rapidly engage cortical resources for balance recovery.^20^ Given evidence showing that balance function and reactive stepping behavior can be improved over repeated exposure of perturbations,^33–35^ future studies could investigate whether behavioral training can modify cortical activity during balance control after stroke and its effect on reactive balance behavior and fall prevention.

### Challenging balance conditions reveal cortical-kinematic relationships during balance recovery in people after stroke

Using smaller perturbation magnitudes in the same cohort reported here, we previously found that delayed cortical N1 responses were associated with slower kinetics of feet-in-place responses only under the most challenging balance condition where laterally directed perturbations shifted body weight onto the paretic leg.^20^ Here, using larger perturbation magnitudes to further increase the challenge, we again found that delayed cortical N1 responses relate to delayed behavioral reactions, now in the form of delayed step initiation. Across studies, brain-behavioral relationships for balance control are consistently strengthened under challenging balance conditions,^20^ suggesting increased reliance on cortical resources for balance control with increasing task difficulty.^11,36^ Given that cortical N1 may reflect cortical error processing for balance control,^16,17^ our results suggest that slowness in cortical error processing may contribute to delayed behavioral reactions under challenging balance conditions after stroke. While we cannot determine causality of cortical activity on reactive balance responses, future studies using neuromodulation techniques to change cortical activity and response latencies may shed light on the causality of temporal features of evoked cortical activity on reactive stepping kinematics.

### Our results indicate that challenging balance conditions effectively index clinically-relevant balance recovery behavior.^4,28,37^

As the participants in this study were instructed to respond using a feet-in-place strategy, stepping responses elicited were unintentional by nature. Compared to volitional steps, unintentional reactive steps have distinct neuromechanical mechanisms that are ecologically relevant for fall prevention.^4,28,37^ For instance, the speed of unintentional reactive steps limits time for motor planning and anticipatory postural adjustment, and are faster and larger than cued, volitional steps.^4,28,37^ Maintenance of upright standing posture under challenging balance conditions may demand greater cortically-mediated processes of response inhibition to suppress the prepotent stepping response, select, and execute alternative strategies.^22^ While speculative, people after stroke may be able to scale up their spatial stepping kinematics to the same degree as controls in unintentional steps elicited under challenging balance conditions, yet still having difficulty speeding up the processes underlying step initiation, partly due to slow central processing and/or response execution. Alternatively, we might be underpowered to detect group-differences in spatial stepping kinematics in the present study.

The cross-sectional nature of this study design precludes determining the casual associations between evoked cortical activity, stepping kinematics, and clinical balance function. Participants in the present study may have developed balance control strategies from the block of smaller perturbations immediately prior to reactive step threshold testing reported here, which might influence balance perturbation tolerance and stepping kinematics. However, it seems unlikely to influence group-level differences given that both groups received the same experimental protocol. While experimenters closely monitored real-time EEG, EMG, and kinetic data to ensure participants returned to a baseline postural state between trials, it is possible that participants were able to predict upcoming perturbations, resulting in subtle changes in anticipatory postural strategies and brain state that have been shown to influence gait and balance in prior studies.^38–40^

## Conclusions

Findings from the present study provide insight into impaired reactive stepping kinematics after stroke under challenging balance conditions and implicate delayed cortical processing. Interventions aiming to facilitate the rapid production of nonparetic stepping responses through accelerating cortical processing may be worth exploring as a potential neurorehabilitation strategy to reduce fall risk for people with stroke.

## Data Availability

All data produced in the present study are available upon reasonable request to the authors

## Funding

This work was supported by the National Institutes of Health [F32HD096816 and K99AG075255] to JP [F32HD105458] to JLM [F32MH129076, Udall Pilot award P50NS098685] to AMP [K12HD055931] to MRB [5T90DA032466, 1P50NS098685, and R01 HD46922] to LHT and [R01 AG072756] to LHT and MRB.

## Notes

### Competing Interest Statement

The authors have declared no competing interest.

### Author Declarations

The experimental protocol was approved by the Emory University Institutional Review Board, and all participants provided written informed consent.

